# Current etiology of hypertension in European children – role of serum uric acid

**DOI:** 10.1101/2024.12.12.24318959

**Authors:** Łukasz Obrycki, Krzysztof Skoczyński, Maksymilian Sikorski, Jan Koziej, Kacper Mitoraj, Jakub Pilip, Michał Pac, Janusz Feber, Mieczysław Litwin

## Abstract

2.

**Background:** While hypertension (HT) in pediatric patients is often secondary (SH), recent trends show a rise in primary hypertension (PH), which is associated with an increasing global prevalence of obesity. Our study aimed to assess the etiology of HT and predictors of PH in a large European cohort of children referred for HT based on office blood pressure (BP) measurements.

**Methods:** We performed retrospective analysis of 2008 children aged 0–18 years (12.3 ± 4.9 years) diagnosed with HT. Patients were classified into white coat hypertension (WCH), PH, or SH groups based on office BP and 24-hour ambulatory BP monitoring (ABPM). Clinical, anthropometric, and biochemical data were collected to differentiate PH and SH and to identify predictors of PH.

**Results:** Out of 2008 patients included in the analysis, HT was confirmed in 1452 patients (556 were classified as WCH). Of 1452 patients with HT: 42.8% had PH, while 57.2% had SH, mainly secondary to renal parenchymal disease (33.2% of SH patients), post-kidney transplant HT (23.1%), aortic coarctation (15.9%) and renovascular HT (13.8%). However, PH started to be the dominant cause of HT after 13 years of age and was diagnosed in 59.1% of 13–18-year-old patients with confirmed HT. Age ≥ 13 years, obesity (BMI-SDS ≥1.65), and serum uric acid ≥ 5.5 mg/dL were identified as significant PH predictors.

**Conclusions:** Our study provides valuable insights into the current etiology of pediatric HT and highlights the role of uric acid level assessment in the diagnosis of PH in children.

## Introduction

Hypertension (HT), one of the main, potentially modifiable cardiovascular risk factors, is a significant clinical problem not only in adult patients but also in children and adolescents. Based on recent studies, it can be estimated that globally HT affects 4–6% of the pediatric population aged 0–18 years, with varying frequency in individual age groups.^1^ In addition to its primary form (primary hypertension, PH), HT is an important complication of other chronic diseases such as diabetes, chronic kidney disease (CKD), vascular pathologies, e.g. coarctation of the aorta, renovascular hypertension, mid-aortic syndrome, and others.^2–9^ However, detailed data on the etiology of HT in the pediatric population is lacking.

Most studies generally show that PH is less common than secondary hypertension (SH)^2–5^ in this age group, but the proportion of PH in pediatric HT patients varied across studies from 15%^2^ to even 46%.^5^ Taking into account an increasing proportion of patients with PH, it is important to distinguish PH from SH at an early stage of the work-up. Data published so far show that at the time of diagnosis, patients with PH are older, overweight, and more often have a family history of hypertension than SH patients.^2,6^ Studies conducted in recent years have shown that patients with PH also have higher serum uric acid concentrations, which may suggest the role of this parameter in the prediction of PH.^10,11^

The aim of our study was to evaluate the etiology of hypertension and predictors of PH in patients diagnosed based on office BP measurements and hospitalized in 2012–2022 at the Department of Nephrology, Kidney Transplantation and Hypertension of the Children’s Memorial Health Institute in Warsaw, Poland.

## Methods

### Study population

A total of 2008 children 0–18 years old (752 girls; mean age 12.3 ± 4.9 years) who had been diagnosed with AH before referral or referred to Children’s Memorial Health Institute in Warsaw (reference hospital for children in Poland with all types of kidney diseases and hypertension) to confirm the diagnosis of AH between January 2012 and December 2022. Our study is a retrospective study, approved by the Bioethics Committee of the Children’s Memorial Health Institute, and was performed in accordance with the Declaration of Helsinki 1975, revised in 2013.

### Data collection

Electronic database was used to identify all patients with confirmed or suspected diagnosis of AH. The initial database query included 2053 patients, 45 of whom were subsequently excluded from the analysis due to lack of data. Patients identified in the electronic database were retrospectively manually reviewed. The date of the initial diagnosis of AH or first attendance in the AH clinic/inpatient ward was identified. Then, patients’ clinical characteristics were collected: age, gender, anthropometric measurements (weight, height, body mass index – BMI, waist circumference), hypertension history, concomitant diseases, laboratory tests (serum creatinine with the calculation of glomerular filtration rate – GFR, serum uric acid level, lipid profile, plasma glucose level, albumin excretion), blood pressure (BP) measurements (office BP, 24-hour ambulatory BP monitoring – ABPM), echocardiography measurements (including left ventricular mass index – LVMi), carotid intima-media thickness (cIMT), pulse wave velocity (PWV) and pulse wave analysis (PWA) including central systolic blood pressure (cSBP).

### Anthropometrics

Height was measured in the upright standing position in older children; in children whose standing height cannot be measured (usually younger than 2 years), height was measured using the portable Harpenden Infantometer and recorded to the nearest 0.5 cm. Body weight was recorded with an accuracy of 0.1 kg using a digital medical scale in participants who were light underwear. Body mass index (BMI) was calculated according to the formula: BMI=body weight (kg)/(height [m])2 and expressed in absolute values and SDS-values based on Polish national normative data.^12,13^ Waist circumference (WC) was measured midway between the lowest rib and the superior border of the iliac crest at the end of a normal expiration with a flexible nonelastic anthropometric tape, to the nearest 0.1 cm and expressed in absolute values and SDS-values based on Polish national normative data.^14^

### Hypertension diagnosis

Patients’ office systolic (SBP) and diastolic (DBP) blood pressure values were measured at least three times during three independent measurements, from which the mean value was calculated. Patients were classified into hypertension stages according to the European Society of Hypertension and Polish Society of Hypertension recommendations,^15,16^ using American reference values (4th Task Force report) for children < 2.5 years old, Polish reference normative values for patients between 2.5 and 16 years old and with standardized cut-off values for older patients.^15–18^ Blood pressure in Table 1 was expressed in absolute values and in SDS-values based on the appropriate normative data^17,18^ for the whole age range to allow comparison between groups.

**Table 1.** Comparison of patients with primary and secondary hypertension.

| Variable | Primary hypertension<br>(n=621) | Secondary hypertension<br>n=(831) | P Value |
| --- | --- | --- | --- |
| Sex | ♀212 (34.1) | ♀353 (42.5) | 0.002 |
|  | ♂409 (65.9) | ♂478 (57.5) |  |
| Age [years] | 15.1 [12.7–16.8] | 10.4 [5.2–14.6] | <0.001 |
| Weight [kg] | 70.5 [55.0–84.0] | 35.9 [19.0–55.0] | <0.001 |
| Height [cm] | 169.0 [158.0–178.0] | 143.5 [114.6–162.0] | <0.001 |
| BMI [kg/m<sup>2</sup>] | 24.0 [21.1–28.3] | 18.1 [15.7–21.3] | <0.001 |
| BMI-SDS | 1.04 [0.16–1.88] | 0.23 [-0.81–1.23] | <0.001 |
| BMI-SDS ≥ 1.04* [№ of patients; (%)] | 127 [45] | 97 [29] | <0.001 |
| BMI-SDS ≥ 1.65† [№ of patients; (%)] | 113 [37] | 30 [6] | <0.001 |
| WC [cm] | 79.5 [71.9–88.0] | 67.0 [60.5–75.5] | <0.001 |
| WC-SDS | 1.23 [0.48–2.05] | 0.45 [-0.44–1.08] | <0.001 |
| BW [kg] | 3.28 [2.83–3.65] | 3.24 [2.80–3.56] | n.s. |
| Gest age [weeks] | 39 [37–40] | 38 [37–40] | 0.038 |
| Apgar score | 10 [9–10] | 10 [9–10] | n.s. |
| SBP [mmHg] | 139 [128–150] | 129 [117–139] | <0.001 |
| SBP-SDS | 2.32 [1.55–2.92] | 2.12 [1.13–3.18] | n.s. |
| DBP [mmHg] | 77 [70–85] | 77 [68–86] | n.s. |
| DBP-SDS | 1.68 [0.84–2.60] | 2.05 [1.02–3.18] | 0.001 |
| 24-h SBP [mmHg] | 124 [117–131] | 128 [119–135] | 0.005 |
| 24-h SBP-SDS | 0.94 [-0.11–1.96] | 1.96 [0.61–2.82] | <0.001 |
| 24-h DBP [mmHg] | 70 [65–74] | 73 [65–80] | 0.01 |
| 24-h DBP-SDS | 0.37 [-0.44–1.22] | 0.77 [-0.39–2.09] | 0.003 |
| 24-h HR [˚/min] | 79 [71–89] | 82 [74–91] | 0.018 |
| cSBP [mmHg] | 119 [112–126] | 117 [107–130] | n.s. |
| HR [˚/min] | 75 [67–83] | 89 [79–102] | <0.001 |
| PWV [m/s] | 5.8 [5.3–6.3] | 5.2 [4.8–6.6] | n.s. |
| PWV-SDS | 1.8 [0.9–2.7] | 1.8 [0.6–3.2] | n.s. |
| PWV-SDS ≥ 1.88 [№ of patients; (%)] | 117 (47.8) | 17 (50.0) | n.s. |
| WCSA [mm<sup>2</sup>] | 7.1 [6.6–7.9] | 7.3 [6.5–8.2] | n.s. |
| cIMT [mm] | 0.43 [0.41–0.46] | 0.44 [0.41–0.49] | 0.001 |
| cIMT-SDS | 0.9 [0.4–1.5] | 1.4 [0.6–2.3] | <0.001 |
| cIMT-SDS ≥ 1.65 [№ of patients; (%)] | 85 ( 20.0) | 78 ( 41.1) | <0.001 |
| LVMi [g/m height<sup>2.7</sup>] | 35.1 [30.4–40.2] | 37.5 [31.6–44.1] | <0.001 |
| LVM/BSA [g/m<sup>2</sup>] | 78.2 [66.1–89.8] | 78.5 [65.0–94.0] | n.s. |
| LVH [№ of patients; (%)] | 73 (16.9) | 62 (26.4) | 0.005 |
| Creatinine [mg/dL] | 0.72 [0.59–0.86] | 0.71 [0.47–1.24] | n.s. |
| GFR [ml/min/1.73 m<sup>2</sup>] | 94.2 [83.5–109.4] | 90.4 [63.4–111.3] | <0.001 |
| Uric acid [mg/dL] | 5.6 [4.8–6.6] | 5.0 [3.9–6.3] | <0.001 |
| Glucose [mg/dL] | 86 [81–91] | 88 [82–96] | <0.001 |
| Insulin [IU/mL] | 13.9 [10.6–19.2] | 14.8 [7.6–21.7] | n.s. |
| **Total cholesterol [mg/dL]** | 159 [137–182] | 168 [147–195] | <0.001 |
| **LDL cholesterol [mg/dL]** | 87 [72–107] | 90 [71–111] | n.s. |
| **HDL cholesterol [mg/dL]** | 50 [42–60] | 54 [45–66] | <0.001 |
| **Triglycerides [mg/dL]** | 85 [65–123] | 93 [66–154] | <0.001 |
| **MALB [mg/24h]** | 8.1 [4.7–14.7] | 10.8 [5.4–34.8] | <0.001 |

\*BMI-SDS $\geq 1.04$ corresponds to the 85<sup>th</sup> percentile (cut-off value for overweight)
†BMI-SDS $\geq 1.65$ corresponds to the 95<sup>th</sup> percentile (cut-off value for obesity)

24-hour ABPM measurements were performed during hospital stay using an oscillometric device (SpaceLabs Monitor 90207) with the most appropriate size of cuff fitted to the non-dominant arm. Readings of SBP, DBP, mean arterial pressure (MAP), pulse pressure (PP), and heart rate (HR) were taken every 20 minutes during the daytime and every 30 minutes at night. ABPM was considered valid if ≥80% of readings were successful. Patients were instructed to complete a diary recording the type and duration of physical activity and sleep period to define daytime/ nighttime periods. Based on collected data, patients were classified into one of the four groups according to recent ESH guidelines (normotension, ambulatory prehypertension, ambulatory hypertension and severe ambulatory hypertension).^15^

### Echocardiography

All echocardiography examinations were performed by examiners in the Echocardiography Laboratory of the Children’s Memorial Health Institute, according to the European Association of Cardiovascular Imaging and American Society of Echocardiography guidelines.^19,20^ Data about left ventricle measurements, relative wall thickness (RWT), and left ventricular mass index (LVMi) were collected. The LVMi was calculated according to de Simone’s formula as a ratio between left ventricular mass and the patient’s height (in meters) to the power of 2.7. To compare results in patients in all age categories, left ventricular hypertrophy (LVH) was defined as an LVMi value ≥95th percentile for age- and sex-based reference data.^21^

### Carotid intima-media thickness and wall cross-sectional area

The cIMT measurements were performed in patients ≥6 years old according to the Mannheim Consensus recommendations using the Aloca Prosound Alpha-7 ultrasound device with a 5.5 to 12.5 MHz linear probe.^22^ The results were given as the mean value of the measurements performed in the left and right common carotid arteries. For analysis of patients in different age categories, both absolute and standard deviation score (SDS) values (obtained by the LMS method) were included. cIMT-SDS ≥1.65 (≥95 percentile) was considered as a cut-off point for increased cIMT (a marker of hypertensive vascular injury).^23–25^

Additionally, the wall cross-sectional area (WCSA) of the common carotid artery was calculated with the use of the following equation:

WCSA = π (dD/2 + cIMT)^2^ - π (dD/2)^2^ [where dD is the mean diastolic diameter of the artery]

### Pulse wave velocity and pulse wave analysis

Data from the oscillometric Vicorder (SMT Medical®) system device were collected, including carotid-femoral pulse wave velocity (PWV) and central systolic blood pressure (cSBP). Measurements were performed according to the published guidelines in the supine position after 5 minutes of rest, at the same time as the BP measurement. Both for PWV and PWA, the first few waves were omitted and, when at least 5 next pulse waves were of good quality, 10–15 consecutive pulse waves (heartbeats) were taken for analysis.^26,27^

The PWV in m/s (absolute values calculated by the device) were subsequently converted to SDS-values based on pediatric normative data.^26–28^ PWV-SDS ≥1.88 (≥97th percentile) were considered as a cut-off point for the increased PWV (a marker of functional hypertensive injury).^26^

cSBP values were interpreted using pediatric normative values obtained with an oscillometric device (Mobil-O-Graph, I.E.M., Stolberg, Germany) and cSBP ≥95^th^ percentile was considered increased.^29^

### Laboratory investigations

Specific biochemical blood serum parameters such as total cholesterol (TC), high-density lipoprotein (HDL), low-density lipoprotein (LDL), triglycerides (TG), glucose, and insulin uric acid (UA) were assessed at the time of diagnosis. In all older patients >6 years old and, when possible, in younger ones, blood samples were taken after 12 hours of fasting. Additionally, the data on 24-hour urinary albumin excretion was collected. Kidney function was assessed using the serum creatinine level measured by the enzymatic method with commercial kits using an autoanalyzer A15 (BioSystems). The estimated glomerular filtration rate (GFR) was calculated with the modified bedside Schwartz formula:^30^

GFR[mL∕min∕1.73m^2^] = 0.413 × H[cm]∕SCr[mg∕dL] H — body length/height

SCr — serum creatinine level

Normal GFR was defined as a GFR value ≥ 90 mL/min/1.73m^2^.

### Classification of patients

All study patients were classified into three groups: white coat hypertension (WCH), primary hypertension, and secondary hypertension. Criteria for the diagnosis of WCH were: (i) hypertension diagnosed based on office BP (all participants), (ii) normotension or ambulatory prehypertension in ABPM measurements, (iii) no pharmacological antihypertensive treatment.

Primary hypertension was diagnosed if: (i) WCH was ruled out, (ii) no secondary causes of hypertension were found, (iii) no medications with the potential to raise BP were administered.

Secondary hypertension (SH) was diagnosed after extensive evaluation of all available data and identification of the cause of hypertension. Those patients were further classified into 13 groups based on SH etiology: 1 – renovascular, 2 – middle-aortic syndrome (MAS), 3 – coarctation of the aorta (CoA), 4 – renal parenchymal, 5 – status post kidney transplantation (KTx), 6 – pheochromocytoma/paraganglioma, 7 – monogenic, 8 – central nervous system (CNS) disorders, 9 – polycystic ovary syndrome (PCOS), 10 – drug-induced, 11 – adrenal diseases, 12 – neuroblastoma, 13 – perinatal history (including prematurity).

### Statistical analyses

All analyzed parameters were checked for normality of distribution with the Shapiro–Wilk test. Because some variables were non-normally distributed, all variables are shown as medians with interquartile ranges and compared using a nonparametric Kruskal–Wallis test. Multivariate regression analysis of clinically important variables was performed to analyze predictors of PH in all patients and patients with normal GFR. P value of <0.05 was considered statistically significant.

## Results

### Classification of patients

Out of 2008 patients (752 girls; 12.3 ± 4.9 years old), after performing ABPM in patients ≥5 years old, WCH was found in 556 subjects (307 girls; 13,7 ± 3.6 years old) and this group was then excluded from further analysis (**Figure 1**). HT was confirmed in 1452 patients (445 girls; 11.8 ± 5.1 years old) and this group was included in the final analysis. The number of patients with confirmed HT increased with patients’ age, with a significant increase from the 14th year of age (**Figure S1**).

**Figure 1.**
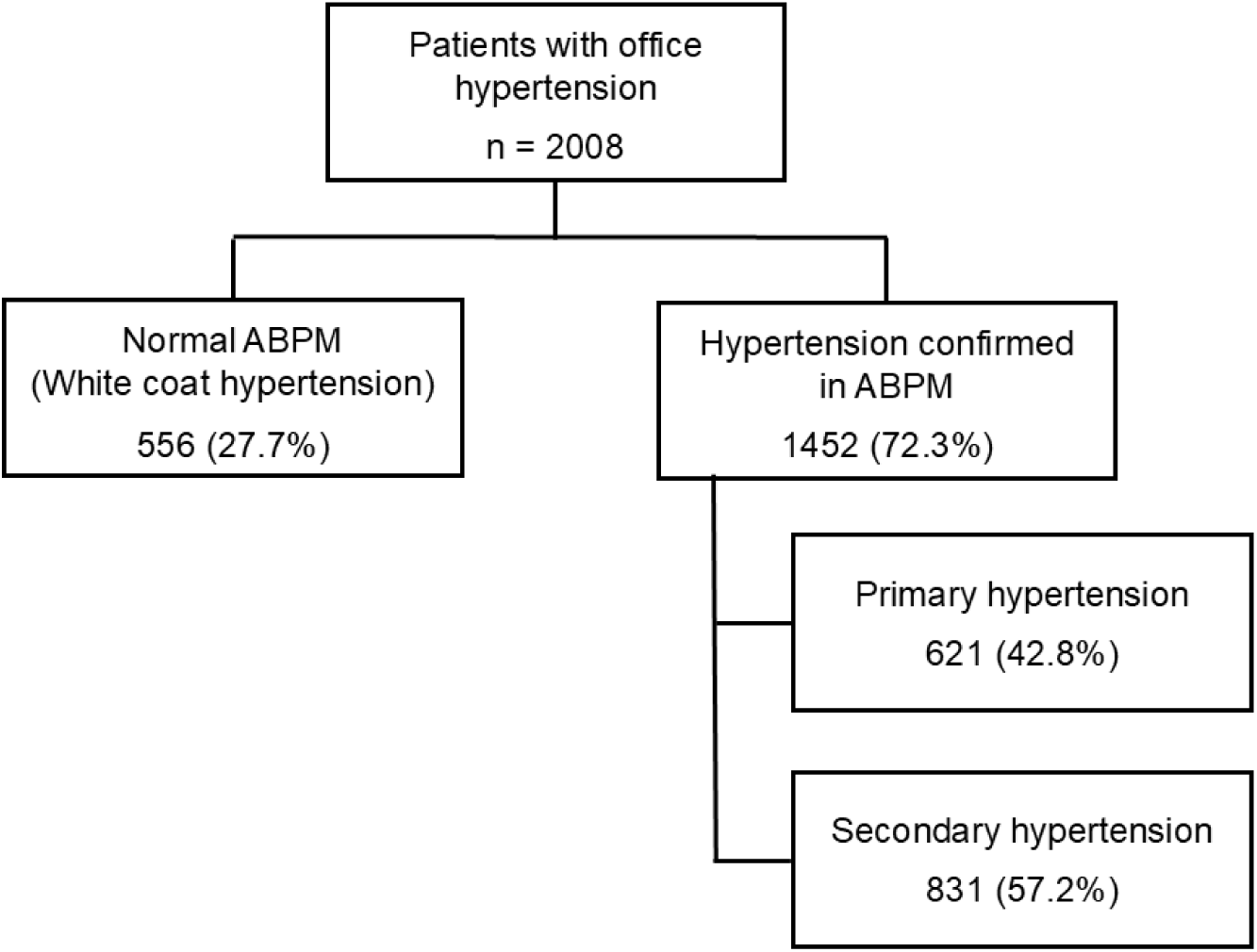
Scheme of the study. ABPM, Ambulatory Blood Pressure Monitoring; HT, hypertension.

After extensive evaluation of all patients, PH was diagnosed in 621 patients (42.8%) and SH in 831 (57.2%) subjects (**Figure 2**). After dividing the patients into age categories, the proportion of patients with PH increased from 33 patients (12.3%) in the group 0–6 years of age; through 105 (28.7%) aged 7–12 years up to 483 (59.1%) patients in the group 13–18 years of age (**Figure 2**).

**Figure 2.**
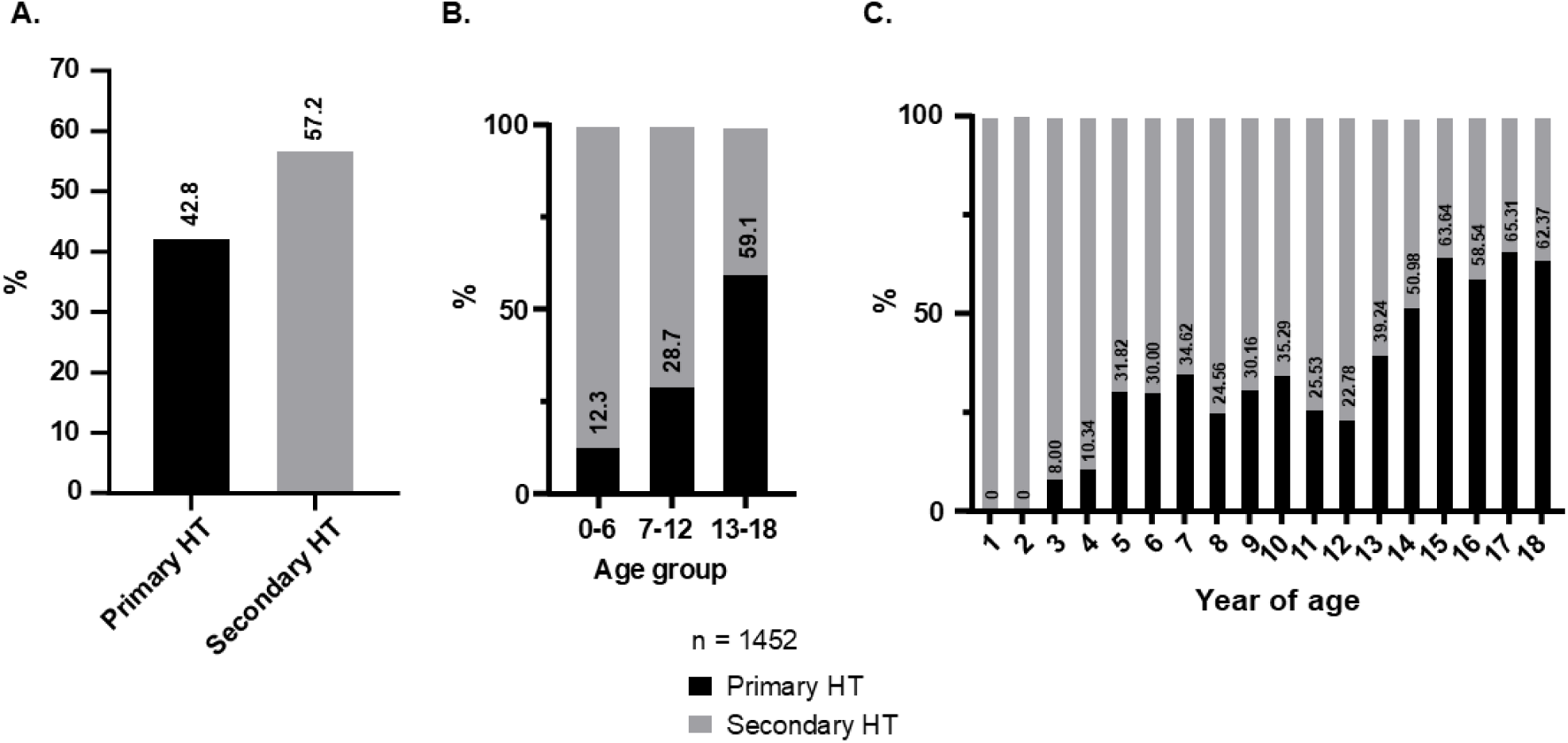
Etiology of hypertension (primary vs secondary) in all patients and in different age categories. Panel. **A** illustrates the etiological profile of HT in the overall study population. **Panel B** presents the categorization of HT etiologies across three distinct pediatric age categories. **Panel C** depicts age-specific distribution of HT etiologies by individual years of age. HT, hypertension.

### Secondary causes of hypertension

Among patients diagnosed with SH, the most common causes were as follows: renal parenchymal disease in 276 patients (33.2%), status post kidney transplantation (KTx) in 192 (23.1%), coarctation of the aorta (CoA) in 132 (15.9%), and renovascular HT in 115 (13.8%) subjects (**Figure 3**). Those four causes were most frequent in all age categories. The only change in their distribution was observed in the group 0–6 years of age, where renovascular cause (15.2%) emerged as the second most common after renal parenchymal etiology (37.7%).

**Figure 3.**
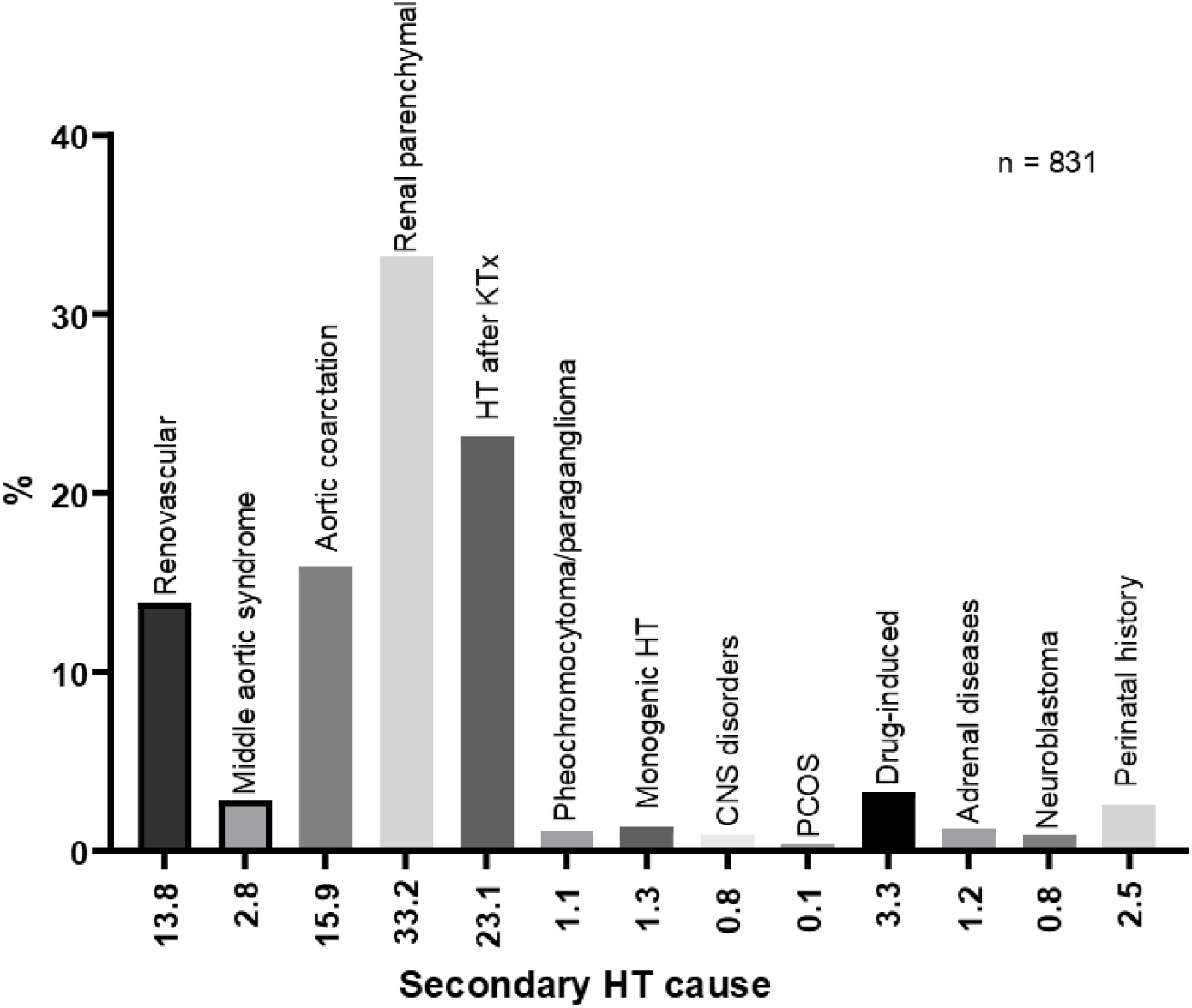
Detailed etiology of hypertension in patients with secondary hypertension. HT, hypertension; KTx, kidney transplantation; CNS, central nervous system; PCOS, polycystic ovary syndrome

Other common conditions responsible for SH were middle-aortic syndrome (MAS) in 23 patients (2.8%), drug-induced hypertension in 27 (3.3%), and perinatal history (including prematurity) in 20 (2.5%). The rest of the causes were observed in 0.1–1.3% of the patients (**Figure 3**).

### Comparison of patients with PH and SH

There were notable anthropometric, hemodynamic, and biochemical differences between patients with PH and SH (**Table 1**). Patients with PH were older (median age 15.1 vs 10.4 years, p<0.001). However, PH was diagnosed as early as 3 years of age in 4 children (8%). All measured anthropometric variables were higher in the PH group, of particular note are measurements related to the risk of overweight/obesity and metabolic syndrome – BMI (24 vs 18.1 kg/m2; p<0.001), BMI-SDS (1.04 vs 0.23; p<0.001), and WC-SDS (1.23 vs 0.45, p<0.001). Consequently, there are significantly more patients with overweight [127 patients (45%) vs 97 (29%); p<0.001] and obesity [113 (37%) vs 30 (6%); p<0.001] in the PH group. There were no significant differences in perinatal parameters (birth weight, gestational age, Apgar score) between groups.

Although office SBP-SDS values did not differ between the PH and SH groups, the SH group showed higher office DBP-SDS values (2.05 vs 1.68, p=0.001). In ABPM, both 24h SBP-SDS (1.96 vs 0.94, p<0.001) and 24h DBP-SDS (0.77 vs 0.37, p=0.003) values were higher in patients with SH.

Consistently, patients with SH presented signs of hypertension-mediated organ damage (HMOD) more frequently: LVMi was higher (37.5 vs 35.1 g/m^2,7^; p<0.001) and LVH was more frequent (26.4% vs 16.9%; p=0.005) in the SH group in comparison with PH group. Patients with SH were also more prone to present signs of vascular damage resulting in higher cIMT (0.44 vs 0.43 mm, p=0.001) and cIMT-SDS values (1.4 vs 0.9, p<0.001). There were no significant differences between groups in PWV, in both absolute and SDS values.

Significant differences were also found between groups in biochemical parameters. The PH group had higher GFR (94.2 vs 90.4 mL/min/1.73m2; p<0.001), higher serum uric acid (5.6 vs 5.0 mg/dL; p<0.001), and lower glucose levels (86 vs 88 mg/dL; p<0.001). A comparison of patients with PH with SH patients and those with normal kidney function (GFR ≥ 90 mL/min/1.73 m^2^) showed even greater differences in serum uric acid concentrations (5.6 vs 4.4 mg/dL; p<0.001). Overall, 91 (56%) of patients with PH had uric acid levels above 5.5 mg/dL compared to 24 (20%) patients with SH (after excluding those with GFR<90 mL/min/1.73m^2^).

Interestingly, patients with PH had significantly different lipid profile: lower total cholesterol (159 vs 168 mg/dL; p<0.001) and triglycerides (85 vs 93 mg/dL; p<0.001), not significantly lower LDL cholesterol (87 vs 90 mg/dL; n.s.), but significantly lower HDL cholesterol (50 vs 54 mg/dL; p<0.001). Patients with SH had significantly higher 24-h microalbumin excretion (10.8 vs 8.1 mg/24h; p<0.001).

### Predictors of primary hypertension

Multivariate regression analysis revealed that the significant predictors of PH were age, BMI-SDS, and serum uric acid level (**Table 2**). Even stronger predictors of PH after exclusion of patients with impaired kidney function (GFR<90 mL/min/1.73m^2^) were age ≥ 13 years; BMI-SDS ≥ 1.65 (≥ 95 percentile; obesity) and uric acid level ≥ 5.5 mg/dL. (**Table 3**).

**Table 2.**
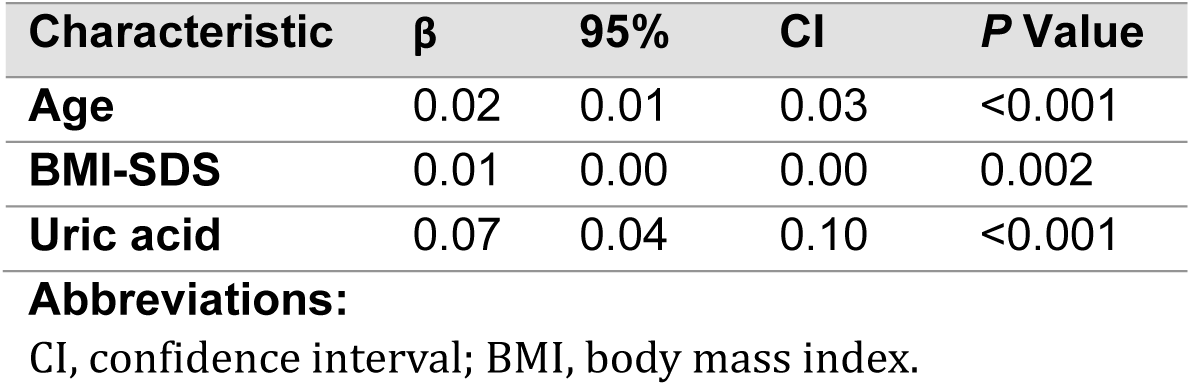
Significant predictors of primary hypertension in all patients.

**Table 3.** Significant predictors of primary hypertension in patients with normal GFR.

| Characteristic | $\beta$ | 95% | CI | *P* Value |
| --- | --- | --- | --- | --- |
| Age ≥ 13 y.o. | 0.15 | 0.07 | 0.23 | <0.001 |
| BMI-SDS ≥ 1.65 | 0.33 | 0.24 | 0.42 | <0.001 |
| Uric acid >5.5 mg/dL | 0.13 | 0.05 | 0.22 | <0.002 |

**Abbreviations:**
CI, confidence interval.

## Discussion

The main outcome of our study is a detailed description of the current (2012–2022) etiology of hypertension in children and adolescents in the largest European cohort of pediatric patients. Another important message is the role of serum uric acid level in predicting PH.

The percentage of patients with primary hypertension in our group of patients (0–18 years) was 42.8%. This is higher than in a study by Chen Y et al. performed in a similar age group (0– 17 years). However, in the study by Chen et al., the BP measurement method was not specified and it is also unclear whether hypertension diagnosed in patients was confirmed by ABPM.^4^ In another study conducted in Turkey (Çakici EK et al.) among 383 participants aged 0–19 years (mean age 13.4 years), the percentage of patients with PH with hypertension confirmed by ABPM was 51.5%.^9^ This difference may be due to the younger age of our patients (11.8 years). Our results compare favorably with a study by Gupta-Malhotra et al., who observed a similar proportion of patients with PH (43%) in a group of 275 hypertensive (office and ABPM) children (3–17 years old).^5^ The results of our study can also be compared to the study by Wyszyńska et al. from 1992, who analyzed the etiology of hypertension in 1025 children referred to our center between 1982 and 1989. It is worth emphasizing that in this historical analysis, no case of PH was found in children under 14 years of age, despite the use of less accurate diagnostic methods. This comparison indicates a significant evolution in the etiology of HT in children and adolescents over the last 3 decades.^31^

### Etiology of secondary hypertension

The most common cause of SH in our study was renal parenchymal HT (33.2%; **Figure 3**), consistent with previously published studies.^4,5,9,31^ Surprisingly, the second most common cause of SH was HT after kidney transplantation (23.1%). However, this is due to the specificity of our center, which is performing approximately 30–50 kidney transplants annually in children and adolescents. In view of the reported prevalence of posttransplant HT ranging between 60% and 90%,^32^ our results showing a large proportion of patients with this etiology of SH, seem justified. The proportion of other causes of SH, such as CoA (15.9%), and renovascular HT (13.8%), are similar to a study by Çakici et al.,^9^ reporting slightly lower percentages of 7% and 9.1%, respectively, and by Wyszynska et al.,^31^ reporting 2.3% (with much greater number diagnosed at Cardiology department) and 9.7%, respectively. In studies by Gupta-Malhotra^5^ et al. and Chen et al.,^4^ the most common causes of SH, after renal parenchymal, were respiratory, rheumatic immune, and hematological diseases. This is most likely also due to the specificity of the centers and the associated available patient pool.

### Comparison of patients with PH and SH

Our cohort demonstrated significant differences in anthropometric measurements, hemodynamic parameters, and biochemical profiles between patients with PH and SH (**Table 1**). Patients with PH were more likely to be male, which is supported by earlier research.^4,5^ They also have significantly higher BMI and BMI-SDS values than patients in the SH group, as already documented in the studies by Chen et al.^4^ and Çakıcı et al.^9^ Our patients with PH also have higher WC and WC-SDS values, as equivalent to abdominal obesity which cannot be compared with other studies because it was not assessed in other studies.

While Gupta-Malhotra et al.^5^ indicated a higher prevalence of preterm birth in the SH patient group, we found no significant differences in terms of gestational age between the PH and SH groups.

The patterns of BP values and HMOD demonstrated significant differences between PH and SH patients. Patients with SH presented significantly higher office DBP-SDS with no differences in office SBP-SDS values. Chen et al.^4^ also found significantly higher office DBP values in the SH group, but in contrast to our study, he also noted significantly higher office SBP values in the PH group. There were no differences in office SBP and DBP values between PH and SH groups in the study by Çakıcı et al.,^9^ office BP was not assessed in the study by Gupta-Malhotra et al.^5^

Patients with SH in our study had significantly higher 24h DBP-SDS values, which is consistent with the study by Çakıcı et al.^9^ Our SH patients also had higher 24h SBP-SDS values, whereas Çakici et al.^9^ found significantly higher 24h SBP-SDS values in the PH group. We cannot compare our ABPM results with studies by Chen et al.^4^ and Gupta-Malhotra et al.^5^ due to the lack of reported ABPM values in these studies.

Our data shows that SH patients have a higher incidence of HMOD and more severe HMOD, including higher LVMi values and a greater frequency of LVH. Additionally, SH patients demonstrated higher cIMT and cIMT-SDS values, indicating a more advanced vascular injury (**Table 1**). These findings are consistent with data reported by Çakıcı et al.,^9^ where the prevalence of LVH was substantial in both PH (36%) and SH patients (27%), and the mean LVMI in all hypertensive patients was 45.28 g/m^2.7^ (compared to 35.1 g/m^2.7^ in PH and 37.5 g/m^2.7^ in SH patients in our groups). However, the study by Çakıcı et al.^9^ identified no significant differences in LVH prevalence or LVMi values between the PH and SH groups.

We noted differences in lipid profile between PH and SH patients: PH patients had significantly lower total cholesterol, triglycerides, LDL cholesterol, and lower HDL cholesterol levels. Differences in the lipid levels between PH and SH patients in our study can be explained by the younger age of the SH cohort, which likely contributes to higher lipid levels, as cholesterol and triglycerides naturally fluctuate during pediatric growth phases. Chen et al. described dyslipidemia (without specifying absolute values) but only in patients with PH, without reflecting on lipid-related factors in patients with SH.^4^

Other biochemical parameters also varied between groups: in SH patients, the 24-hour microalbumin excretion was higher, corresponding to findings by Gupta-Malhotra et al.^5^ The SH group also demonstrated higher glucose levels (**Table 1**).

Additionally, corticosteroids often used as an immunosuppressive treatment after kidney transplantation, are known to increase lipid levels and insulin resistance, and influence glucose metabolism. Finally, blood samples in the youngest group (mostly with SH) were taken without fasting, which potentially could affect serum lipid and glucose results.

### Serum uric acid levels

PH patients had significantly higher uric acid levels compared to those with SH (**Table 1**), which is in agreement with a previous paper by Feig et al. who compared groups with different blood pressure status and with PH and SH.^10,11^ In his study, serum uric acid >5.5 mg/dL was found in 89% of subjects with PH compared to 30% with SH, 0% with white-coat hypertension, and 0% of the control normotensive group. In our study serum uric acid >5.5 mg/dL was found in 56% of patients with PH and 20% of those with SH.

### Predictors of PH – multivariate analysis

In a multivariate regression analysis, older age, and BMI-SDS emerged as strong predictors of PH. WC-SDS was not found to be a significant predictor of PH in our analysis when analyzed simultaneously with BMI, most likely due to strong cross-correlation between BMI and WC. Our findings are in agreement with the conclusions previously described by various authors, who also identified age and BMI as key PH indicators, often accompanied by a family history of hypertension.^4,5,9^ Another significant predictor of PH was serum uric acid (**Table 2**). Moreover, after analyzing PH predictors only in patients with normal kidney function, we found that uric acid level > 5.5 mg/dL was a significant predictor of PH (**Table 3**). Our finding is consistent with studies indicating that increased UA may contribute to early cardiovascular changes by inducing endothelial dysfunction and impairing nitric oxide bioavailability, ultimately leading to elevated blood pressure.^33^ Higher UA levels are perceived as one of the non-classical components of metabolic syndrome and have been associated with greater cardiovascular risk, even in young populations.^33–36^

## Perspectives

Our study, including the largest European cohort of children with hypertension described in recent years, provides detailed information on the current etiology of hypertension in children and adolescents and its evolution over the last decades. We confirmed the role of anthropometric parameter assessment in diagnosing PH, but we also showed, based on multivariate regression analysis, that serum uric acid plays a major role in predicting PH in children and adolescents. Future research could expand on these predictors to develop a robust, risk-based model for pediatric hypertension screening that includes genetic, metabolic, and lifestyle factors. Such a model would enable more personalized treatment strategies, especially in populations experiencing increasing rates of pediatric obesity and metabolic syndrome. Our study also sets the stage for longitudinal studies to assess the long-term impact of early PH management, particularly in reducing adult cardiovascular risks, e.g. by reducing serum acid levels non-pharmacologically and, when needed, with pharmacotherapy.

## 6. What is new?

Our study:

- Highlights the rising prevalence of primary hypertension in children;
- Provides detailed comparative data on clinical characteristics between primary and secondary hypertension in young patients;
- Identifies age, BMI-SDS, and serum uric acid level as key predictors of PH in children.

## 7. What is relevant?

- Serum uric acid level should play an important role in differentiating the etiology of hypertension and assessing cardiovascular risk in pediatric patients with elevated blood pressure.
- The increasing rate of PH among the youngest children emphasizes the need to take into account the diagnosis of PH also in this age group.

## 8. Clinical/Pathophysiological implications

- Awareness of the increasing incidence of PH in the pediatric population should result in greater efforts to eliminate PH risk factors such as overweight and elevated uric acid levels very early.
- Analysis of hemodynamic phenotype may help in diagnosing the etiology of HT

## Data Availability

Data available on request from the authors

